# Perceived comfort with migraine treatments and control of migraine symptoms: results from the HEAD-US study

**DOI:** 10.64898/2026.01.24.26344649

**Authors:** Kevin M. Spiegler, Ali Ezzati, Kristina M. Fanning, Ryan Bostic, Alexandre Urani, François Cadiou, Richard B. Lipton, Angeliki Vgontzas

## Abstract

**Objective:** To identify patient characteristics and medication factors associated with perceived comfort with migraine treatments and control of migraine symptoms.

**Background:** Patient perceptions of migraine treatment influence adherence and outcomes, yet little is known about their underlying drivers. Our study objective was to identify clinical features of migraine associated with comfort and perceived control of symptoms, and medication classes associated with higher patient comfort and efficacy.

**Methods:** Participants in the Headache Assessment via Digital Platform in United States (HeAD-US) study completed a cross-sectional survey on demographics, migraine burden, mood, disability, and medication use. Comfort and control were defined using corresponding items from the Migraine Treatment Optimization Questionnaire (mTOQ-6) and categorized as “high” or “low”. We compared patient characteristics, clinical features, and medication classes by reported comfort and control.

**Results:** Among 5717 participants (mean age 41.5 ± 13.1 years, 91.2% female), 58.5% reported high comfort with migraine treatments, while only 27.5% reported high control of migraine symptoms. High comfort was associated with fewer monthly headache days (mean 9.9 vs. 14.2, p<0.001), lower migraine symptom severity (median MSSS 18 vs. 19, p<0.001), and lower disability (MIDAS severe: 72.7% vs 91.2%, p<0.001) and mood symptom scores (PHQ-4 severe: 9.5% vs. 21.0%, p<0.001). High perceived control showed similar associations with employment (OR=1.25, CI 1.15-1.48, p = 0.013), monthly headache days (mean 9.0 vs 12.7,p<0.001), migraine severity (median MSSS 18 vs 19, p<0.0001), disability (severe: 66.0 vs 85.8, p<0.001), and mood symptoms (severe: 7.6 vs 16.9, p<0.001) but was not associated with income or education. Among those on monotherapy, gepants and triptans were associated with higher comfort and efficacy than OTC medications or opioids/barbiturates. For preventive therapy, beta blockers and botulinum toxin were associated with the lowest perceived comfort and efficacy.

**Conclusion:** Perceived comfort and control were linked to headache frequency, severity and disability and mood symptoms. Medication class use influenced perceptions, with gepants and triptans rated most favorably. These findings underscore the importance of incorporating patient perspectives into treatment planning, with particular attention to mood, disability, and choice of medication.

## Introduction

Patient perception of illness plays a critical role in health outcomes, influencing treatment adherence, symptom reporting, and healthcare utilization across a range of chronic conditions, including migraine.^1–4^ Disease perceptions may shape how patients manage symptoms and engage with care, but the factors that influence such perceptions remain poorly defined.

In migraine, patient-reported outcomes (PROs) have become increasingly important for capturing the lived experience of disease. PROs assess symptoms, functioning, and health perceptions directly from patients, complementing clinical measures such as attack frequency or biomarkers.^5,6^ These patient-centered measures are especially relevant in migraine, where treatment goals extend beyond reducing attack frequency to improving daily functioning and quality of life. Incorporating PROs into routine care can provide clinicians with a more complete view of patient well-being and guide treatment choices that align with patient preferences.

Patient perceptions of treatment effectiveness and symptom control play a crucial role in migraine management, influencing both treatment adherence and reported symptom severity.^7–9^ Prior studies indicate that patients value complete and rapid relief from abortive migraine medication.^10,11^ On the other hand, inconsistent acute treatment response increases the risk of progression to chronic migraine.^12^ However, limited data exist on how patients’ perceived comfort with treatment and perceived efficacy of medications relate to headache severity, mood, and disability. This is critical, given evidence that perceived disability and actual disability can diverge, with implications for healthcare utilization and outcomes.^13–17^

Patient comfort with their medication strategy is vital for effective management. Studies have shown that negative beliefs regarding medical interventions for migraine lead to poorer outcomes and increased report of adverse events.^18–20^ Similarly, a diminished sense of control over symptoms is associated with greater disability, psychological stress, and migraine chronification.^12,21^ Self-efficacy, or the perceived ability to control one’s migraine, has been linked to better adherence and reduced disability. In headache populations, lower self-efficacy is identified as a barrier to treatment uptake and is associated with greater headache impact, while intervention studies increasing self-efficacy report improvements in disability.^22^

The Headache Assessment via Digital Platform in United States (HeAD-US) study has previously described migraine treatment patterns and diagnostic validity using real-world data collected via a smartphone app.^23^ A recent study from this cohort described efficacy of acute medications. ^24^ In this study, we extend this work by examining how patients perceive comfort with migraine medications and control of migraine symptoms, and how these perceptions are shaped by demographics, clinical features, and medication class. We hypothesize that lower comfort and control reflect more severe disease burden and are influenced by both patient factors and the types of medications used.

## Methods

### Study design and participants

The HeAD-US study is a digital cohort study of adults in the United States with migraine, recruited via the Migraine Buddy smartphone application, which is a commercially available and used by individuals with headaches to track their symptoms. Detailed methodology of the HeAD-US study has previously been published.^24–26^ In brief, between September and December 2023, app users were invited to participate in the HeAD-US study via in-app notifications. Participation was voluntary, and no financial compensation or incentives were offered. Participants provided electronic consent and subsequently received the study questionnaire through the app. All study procedures were approved by the Institutional Review Boards at the University of California Irvine (CIRBI protocol # Pro00072897) and at Brigham and Women’s Hospital (IRB # 2025P000071).

Participants were eligible for the study if they were aged 18 years or older and provided consent. Participants were excluded if they did not meet International Classification of Headache Disorders, 3rd edition (ICHD-3) criteria for migraine as assessed through the American Migraine Study (AMS)/American Migraine Prevalence and Prevention (AMPP) diagnostic module or if they did not complete the two items on perceived comfort with medications and control over symptoms after taking medication from the Migraine Treatment Optimization Questionnaire (mTOQ-6).^27,28^

### Study Measures

#### Independent variables

The sample of participants were defined twice, based on high or low levels of comfort with treatment and perceived control over symptoms. Comfort was defined using item 5 from the mTOQ-6 (“Are you comfortable enough with your migraine medication to be able to plan your daily activities?”) and control was defined using item 6 (“After taking your migraine medication, do you feel in control of your migraines enough so that you feel there will be no disruption to your daily activities?”).

These items were specifically chosen for their subjective nature, whereas mTOQ-6 items 1-4 require more objective responses regarding medication efficacy. It should be noted that each item of the mTOQ-6 measures a distinct domain of treatment response and each individual item had meaningful factor loading at validation.^28^ Patients responded to each question using a 4-point Likert scale with options of “never”, “rarely,” “less than half of the time,” or “more than half of the time.” To improve interpretability of outcomes, responses were dichotomized into “*low*” (never/rarely/less than half) and “*high*” (more than half the time) categories for both comfort and control.

#### Outcome measures

The primary outcomes were monthly headache days, symptom severity, reported disability, and mood symptomatology. The following tools were used to measure these:

##### Headache frequency

Patients were asked to report their average number of headache days per month.

##### Migraine Symptom Severity Score (MSSS)

The MSSS is a validated tool utilized to measure the frequency of seven ICHD-3 diagnostic migraine symptoms (unilateral pain, pulsatile pain, moderate or severe pain intensity, provocation with exertion, photophobia, phonophobia and nausea).^29^ Responses for each item range from 0-3 (never, rarely, less than half of the time or more than half of the time), with a maximum score of 21.

##### Migraine Disability Assessment Score (MIDAS)

The MIDAS questionnaire measures headache-related disability^30^, defined as the number of days impacted by migraine over the last 3 months across 3 domains: work, chores, and non-work activities. The total MIDAS score was utilized, with 0-5 are considered little to no disability, 6-10 considered mild, 11-20 considered moderate, and 21 or more considered severe disability.

##### Patient Health Questionnaire-4 (PHQ-4)

The PHQ-4 is a 4-item survey designed to screen for anxiety and depression symptoms in clinical settings.^31^ Total scores range from 0-12, with 0-2 considered normal, 3-5 as mild, 6-8 as moderate, and 9-12 as severe mood symptoms.

Participants also reported current use of acute medication classes (Over the counter (OTC), triptan, gepant, ergot, barbiturate, opioid) and preventive medications (antidepressant, anti-seizure, beta blocker, OnabotulinumtoxinA, gepant, CGRP monoclonal antibody). Multiple selections were allowed, and participants could respond “unsure” if medication class was unclear.

Lastly, participants were asked to report on demographic characteristics (age, biological sex, race, ethnicity, annual household income, employment status, and level of education).

### Statistical Analysis

All statistics were run using IBM Statistical Package for Social Science (SPSS) version 28.^32^ All variables were assessed for collinearity where appropriate. Comparisons were run across both comfort and control measures against demographic data and outcome measures individually. Normality was assumed given large sample sizes. Student’s t-test was used for numerical data, median test for ordinal data, and Chi square test for categorical data. Alpha was set to 0.05 for all evaluations. Levene’s test was used to test for variance where appropriate. Bonferroni correction was used for all post-hoc testing. Missing or optional non-responses were treated as missing data in all comparisons. Subsequently, logistic regression was used to identify predictive variables for comfort and control variables. Only items reaching statistical significance in the initial comparisons were included in the regressions.

Multivariable logistic regressions were then performed to identify independent predictors of high comfort and high control. Only variables that were significant in univariate analyses were included in the regression models.

To examine the relationship between medication use and patient perceptions, we conducted subgroup analyses:

- *Number of medications:* Comfort and control were compared across groups reporting 0, 1, or ≥2 acute or preventive medications.
- *Medication class (monotherapy only):* Among participants on a single acute or preventive medication, logistic regressions were conducted to compare comfort and control across classes. Pairwise comparisons rotated the reference group to evaluate relative differences. Due to small sample sizes, ergot, barbiturate, and opioid users were pooled into a single group.

## Results

### Sample Overview

A total of 6810 participants were eligible for the study. Of these, 543 did not meet IHCD-3 criteria for a diagnosis of migraine and were excluded. An additional 550 did not complete items 5 and 6 of the mTOQ-6 and were also excluded. The final study cohort included 5717 participants with a mean age of 41.5 years (SD = 13.1), 91.5% female and 88.3% white. In the overall sample, 3345 (58.5%) reported high comfort, whereas 1575 (27.5%) reported high control.

Participants reporting high comfort were significantly older (Table 1A, mean age: 42.4 vs. 40.7 years, *p* < 0.001), more likely to be employed (74.8% vs. 62.9%, *p* < 0.001), and had higher income and education levels (≥$75,000: 57.9% vs. 44.2%, *p* < 0.001; ≥12 years education: 96.7% vs. 94.1%, *p* < 0.001). They were also less likely to report Hispanic ethnicity (5.79% vs 7.40%, *p* = 0.015). There were no significant differences in sex or race.

**Table 1A:**
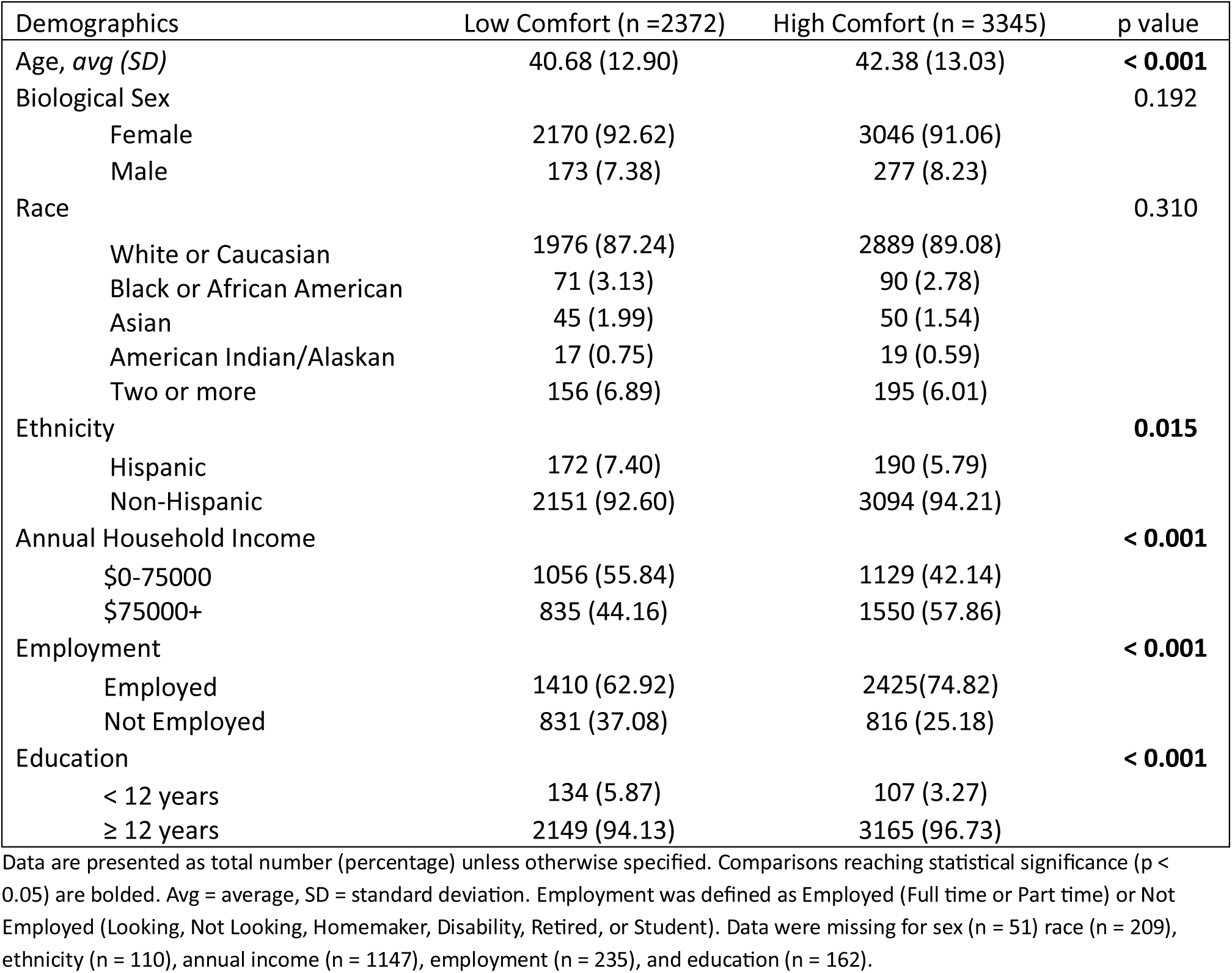
Demographic characteristics in the HeAD-US study by level of comfort with migraine treatments.

Participants with high control were also older (Table 1B, mean age: 43.9 vs. 40.8 years, *p* < 0.001) and more likely to be employed (74.6% vs. 68.2%, *p* < 0.001). Income and education were modestly higher but not significant in multivariable analysis. No significant differences were observed by sex, race, or ethnicity.

**Table 1B:** Demographic characteristics in the HeAD-US study by level of control of migraine symptoms.

| Demographics | Low Control (n = 4142) | High Control (n = 1575) | p value |
| --- | --- | --- | --- |
| Age, *avg (SD)* | 40.82 (12.76) | 43.93 (13.39) | **< 0.001** |
| Sex |  |  | 0.163 |
| Female | 3788 (92.37) | 1428 (91.25) |  |
| Male | 313 (7.63) | 137 (8.75) |  |
| Race |  |  | 0.899 |
| White or Caucasian | 3519 (88.28) | 1346 (88.44) |  |
| Black or African American | 112 (2.81) | 49 (3.22) |  |
| Asian | 70 (1.76) | 25 (1.64) |  |
| American Indian/Alaskan | 27 (0.68) | 9 (0.59) |  |
| Two or more | 258 (6.47) | 93 (6.11) |  |
| Ethnicity |  |  | 0.328 |
| Hispanic | 255 (6.26) | 107 (6.98) |  |
| Non-Hispanic | 3819 (93.74) | 1426 (93.02) |  |
| Annual Household Income |  |  | **< 0.001** |
| \$0-75000 | 1671 (50.20) | 514 (41.42) | |
| \$75000+ | 1658 (49.80) | 727 (58.58) | |
| Employment |  |  | **< 0.001** |
| Working | 2697 (68.16) | 1138 (74.62) |  |
| Not Working | 1260 (31.84) | 387 (25.38) |  |
| Education |  |  | **0.003** |
| < 12 years | 195 (4.85) | 46 (3.00) |  |
| ≥ 12 years | 3827 (95.15) | 1487 (97.00) |  |

Data are presented as total number (percentage) unless otherwise specified. Comparisons reaching statistical significance ( $p < 0.05$ ) are bolded. Avg = average, SD = standard deviation. Employment was defined as Employed (Full time or Part time) or Not Employed (Looking, Not Looking, Homemaker, Disability, Retired, or Student). Data were missing for sex (n = 51) race (n = 209), ethnicity (n = 110), annual income (n = 1147), employment (n = 235), and education (n = 162).

### Associations Between Comfort/Control and Clinical Severity

#### Comfort

Compared to those with low comfort, those with high comfort reported on average 4.31 less monthly headache days (Figure 1A, t(4571.175) = 19.922, p < 0.001), lower MSSS scores (Figure 1C, χ²(1) = 91.735, p <0.001) and lower MIDAS scores (Figure 2A, χ²(3) = 306.603, p < 0.001) with post hoc testing significant at all levels. Lower PHQ-4 scores also were associated with higher reported comfort (Figure 2C, χ²(1), = 216.010, p < 0.001). Additional data is provided in Supplemental Table 1A.

**Figure 1.**
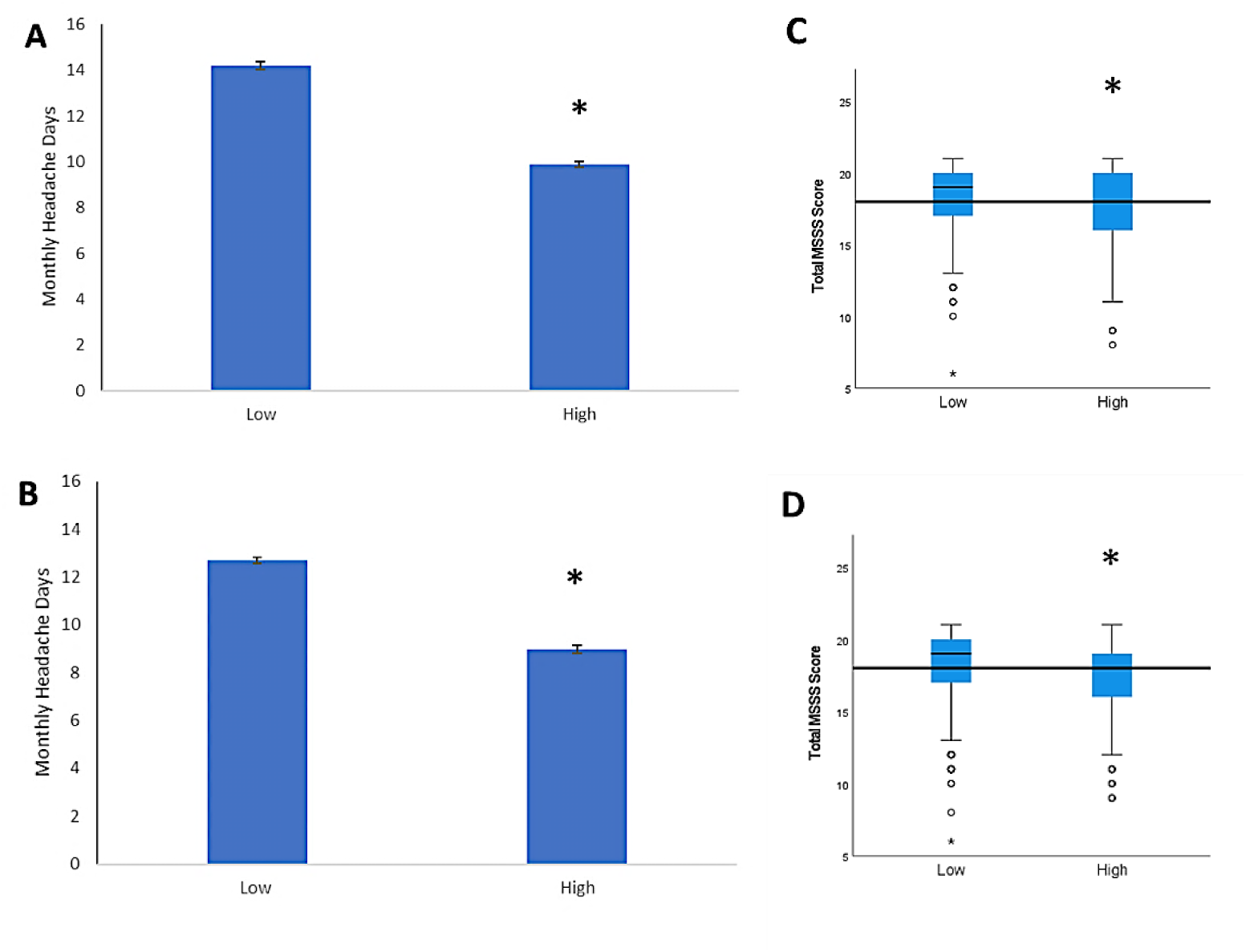
Monthly Headache Days and Migraine Symptom Severity Score. Figure (A) depicts average monthly headache days across high (n = 3345) and low (n = 2372) reported comfort. Figure (B) shows the same across high (n = 1575) and low (n = 4142) reported control. Box plots show total MSSS score across reported low and high comfort (C) and control (D). The horizontal line highlights median of the whole cohort. Error bars indicate standard error. * denotes statistical significance (p < 0.05).

**Figure 2.**
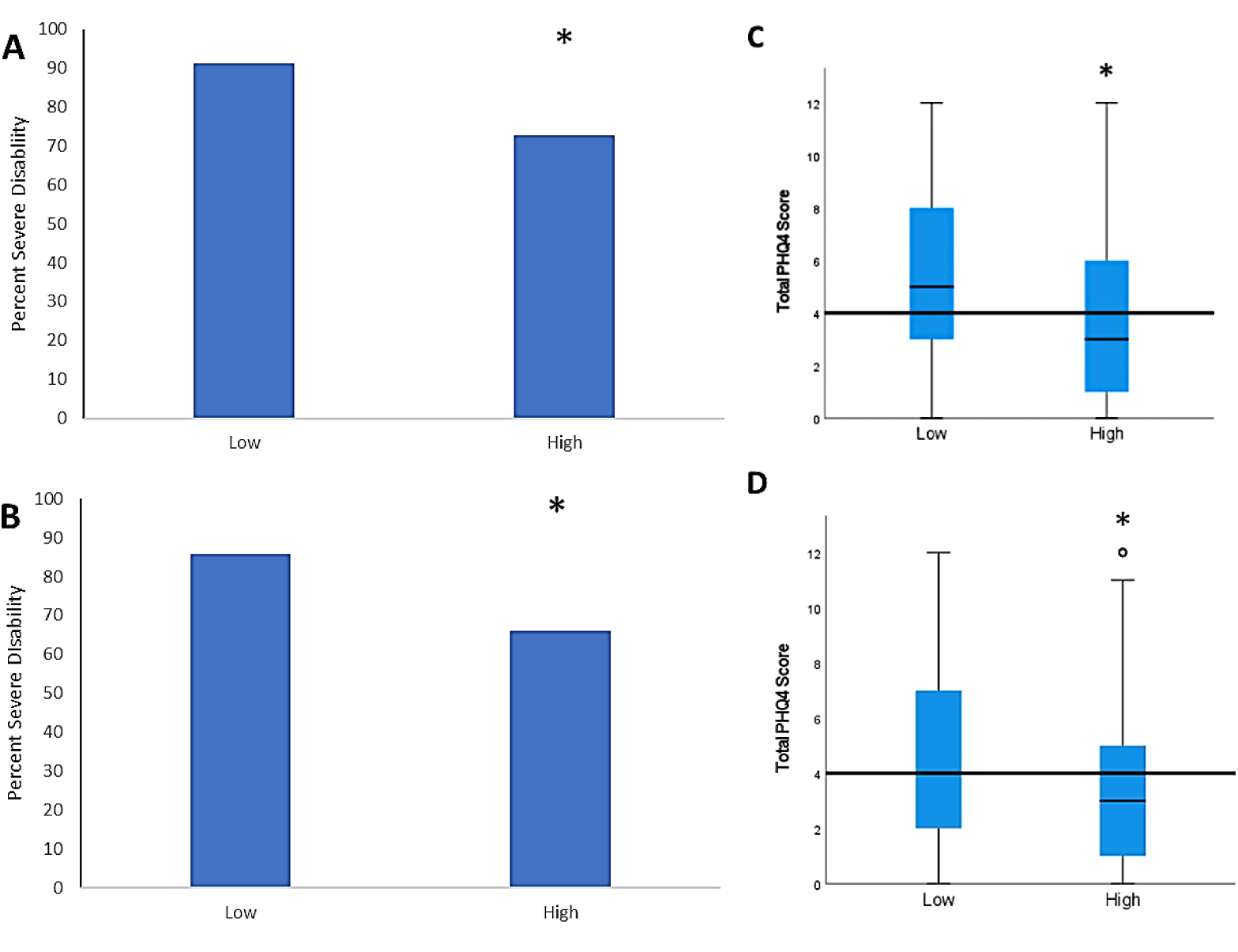
Migraine Disability Assessment Score and Patient Health Questionnaire-4. Figure (A) depicts percentage of participants indicating severe disability based on MIDAS score across high (n = 3345) and low (n = 2372) reported comfort. Figure (B) shows the same across high (n = 1575) and low (n = 4142) reported control. Box plots show total PHQ-4 score across reported low and high comfort (C) and control (D). The horizontal line highlights median of the whole cohort. Error bars indicate standard error. * denotes statistical significance (p < 0.05).

#### Control

Similarly, those with high control reported on average 3.69 less monthly headache days (Figure 1B, t(3473.052) = 17.270, p < 0.001). In line with this, MSSS scores were lower amongst those with high reported control (Figure 1D, χ²(1) = 72.317, p < 0.001). Relatively lower MIDAS scores were associated with higher control (Figure 2B, χ²(3) = 308.429, p < 0.001) with post hoc testing significant at all levels. PHQ-4 scores were lower in those with high control as well (Figure 2D, χ²(1) = 149.781, p < 0.001). Additional data is provided in Supplemental Table 1B.

### Multivariable Predictors of Comfort and Control

#### Comfort

Greater monthly headache frequency (OR = 0.962, *p* < 0.001) and higher MSSS scores (OR = 0.929, *p* < 0.001) were significantly associated with lower comfort (see Table 2A). Similarly, participants with moderate (OR = 0.524, *p* < 0.001) and severe (OR = 0.261, *p* < 0.001) disability on the MIDAS scale were less likely to report high comfort, compared to those with little or no disability (*p* < 0.001) as were those with mood symptoms on the PHQ-4 (*p* < 0.001). Other significant associations with high comfort included older age (OR 1.006, p = 0.047), higher annual income (OR 1.299, p < 0.001), education level (OR 1.506, p = 0.013) and current employment (OR 1.499, p < 0.001). Ethnicity was not associated with comfort level (p = 0.157).

**Table 2A:** Logistic Regression for prediction of high comfort response.

| Variable | Odds Ratio | 95% CI | p value |
| --- | --- | --- | --- |
| Age | 1.006 | 1.000-1.011 | **0.047** |
| Ethnicity (Ref: Not Hispanic) | 0.829 | 0.639-1.075 | 0.157 |
| Annual Income (Ref: <\$75,000) | 1.299 | 1.137-1.485 | **<0.001** |
| Education (Ref: <12 years) | 1.506 | 1.090-2.081 | **0.013** |
| Employment (Ref: Not Working) | 1.499 | 1.292-1.742 | **<0.001** |
| Monthly Headache Days | 0.962 | 0.954-0.970 | **<0.001** |
| MSSS | 0.929 | 0.901-0.958 | **<0.001** |
| PHQ4 (Ref: Normal) |  |  | **<0.001** |
| Mild | 0.642 | 0.545-0.756 | *** |
| Moderate | 0.529 | 0.440-0.637 | *** |
| Severe | 0.368 | 0.298-0.455 | *** |
| MIDAS (Ref: Little to None) |  |  | **<0.001** |
| Mild | 0.816 | 0.420-1.586 |  |
| Moderate | 0.524 | 0.295-0.931 | *** |
| Severe | 0.261 | 0.152-0.449 | *** |

Comparisons reaching statistical significance ( $p < 0.05$ ) are bolded. \*\*\* denotes significant subcategories for categorical variables. *Ref* indicates reference group for the categorical variable. MSSS = Migraine Symptoms Severity Score, PHQ4 = Patient Health Questionnaire-4, MIDAS= Migraine Disability Assessment Score

#### Control

As with comfort, greater headache frequency (OR = 0.962, p < 0.001) and higher MSSS scores (OR = 0.947, p < 0.001) were associated with lower control, though changes in odds per unit increase were modest (see Table 2B). Participants with increasing levels of mood symptoms reported on PHQ-4 again had lower odds of high control (*p* < 0.001) as did increasing disability on MIDAS (*p* < 0.001). Older age (OR = 1.016, p < 0.001) and current employment (OR = 1.245, p = 0.013) were positively associated with perceived control. Income and education level were not significant predictors in the final model (p = 0.538 and 0.152, respectively).

**Table 2B:** Logistic Regression for prediction of high control response.

| Variable | Odds Ratio | 95% CI | p value |
| --- | --- | --- | --- |
| Age | 1.016 | 1.010-1.022 | **<0.001** |
| Annual Income (Ref: <\$75,000) | 1.047 | 0.904-1.212 | 0.538 |
| Education (Ref: <12 years) | 1.342 | 0.897-2.007 | 0.152 |
| Employment (Ref: Not Working) | 1.245 | 1.047-1.481 | **0.013** |
| Monthly Headache Days | 0.962 | 0.952-0.972 | **<0.001** |
| MSSS | 0.947 | 0.918-0.978 | **<0.001** |
| PHQ4 (Ref: Normal) |  |  | **<0.001** |
| Mild | 0.637 | 0.539-0.752 | *** |
| Moderate | 0.619 | 0.506-0.756 | *** |
| Severe | 0.351 | 0.269-0.460 | *** |
| MIDAS (Ref: Little to None) |  |  | **<0.001** |
| Mild | 0.791 | 0.507-1.234 |  |
| Moderate | 0.554 | 0.375-0.820 | *** |
| Severe | 0.313 | 0.218-0.451 | *** |

Comparisons reaching statistical significance ( $p < 0.05$ ) are bolded. \*\*\* denotes significant subcategories for categorical variables. *Ref* indicates reference group for the categorical variable. MSSS = Migraine Symptoms Severity Score, PHQ4 = Patient Health Questionnaire-4, MIDAS= Migraine Disability Assessment Score

### Impact of Medication Use and Class on Comfort and Control

#### Acute medications

Among the 5717 participants, 370 (6.5%) reported no rescue medications, 2420 (42.3%) were taking one rescue medication, and 2927 (51.2%) were taking two or more. Those taking no rescue medication reported lower comfort levels than those taking either one rescue medication or two or more (Figure 3A, p < 0.001). Regarding perceived control, participants using one rescue medication were more likely to report high control than those taking either no rescue medication or two or more (Figure 3B, p < 0.001).

**Figure 3.**
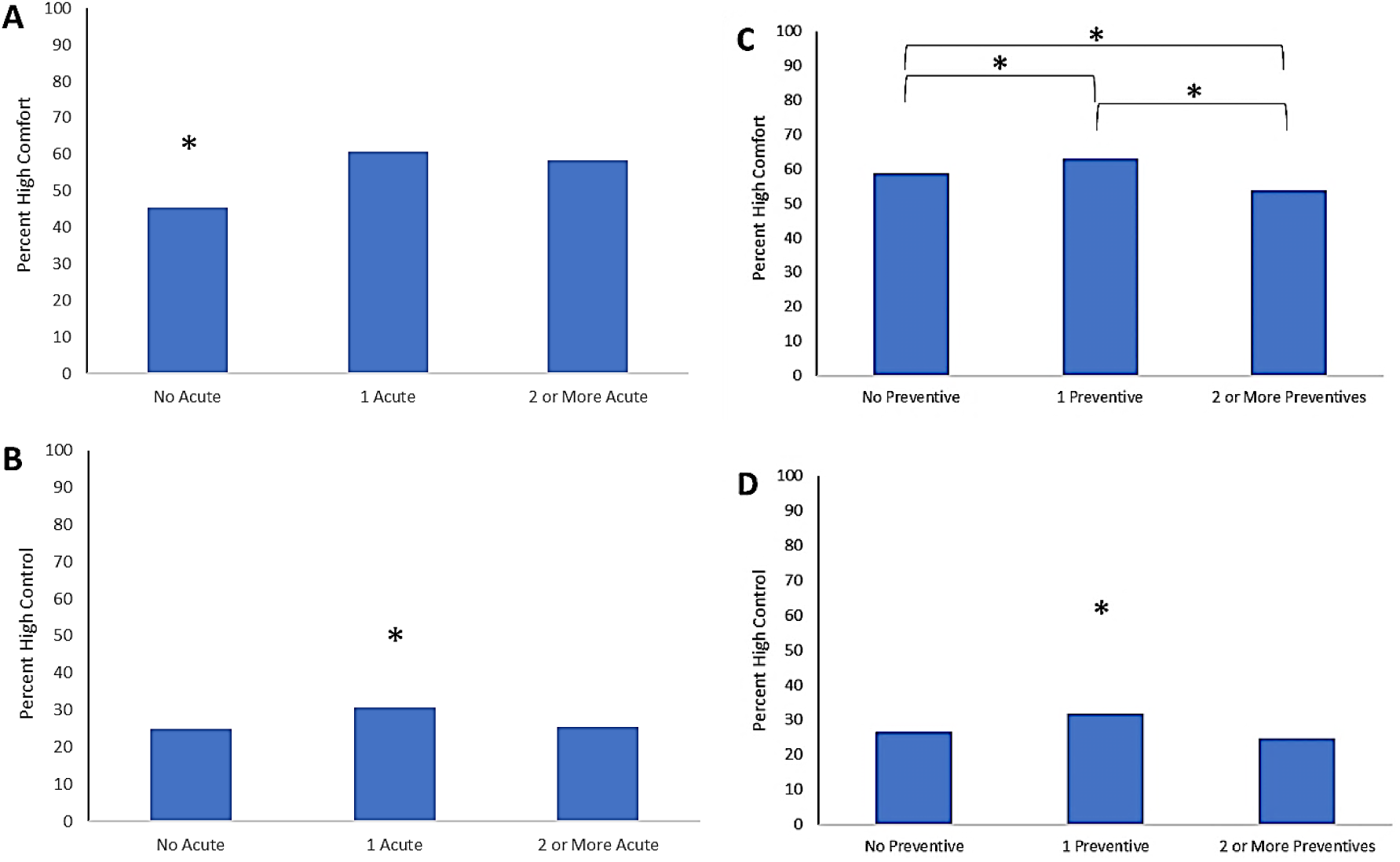
Number of Medications. Figure A shows the percentage of participants reporting high comfort taking no rescue medication (n = 370), one rescue medication (n = 2420), or two or more rescue medications (n = 2927). Figure B displays percentage reporting high control across the number of rescue medications. Figure C depicts the percentage of participants reporting high comfort taking no preventive medication (n = 2135), one preventive medication (n = 1751), or two or more preventive medications (n = 1831). Figure D again shows similar data for the control variable. * denotes statistical significance after Bonferroni correction (p < 0. 017).

#### Preventive Medications

Of the total sample, 2135 (37.3%) reported using no preventive medication, 1751 (30.6%) reported using one preventive medication, and 1831 (32.0%) reported using two or more. Regarding comfort, those taking one preventive medication were most likely to report high comfort, followed by those using two or more preventives, with those without a preventive medication reporting the least comfort (Figure 3C, p < 0.001). Lastly, participants with 1 preventive medication reported high control more frequently relative to the other groups (Figure 3D, p < 0.001).

### Comfort and Control by Acute Medication Class

Among the 2420 patients using only one rescue medication, 1018 (42.1%) used a triptan, 727 (30.1%) used an OTC medication, 570 (23.6%) used a gepant, and 105 (4.3%) used an ergot, barbiturate, or opioid.

#### Comfort

Compared to participants in the ergot/barbiturate/opioid monotherapy group, participants using only triptans (p = 0.004) or gepants (p = 0.003) were significantly more likely to report high comfort (Table 3A). No difference was observed between OTCs and the ergot/barbiturate/opioid group (*p* = 0.672). High comfort rates did not differ between triptans and gepants (p = 0.672), but both were rated significantly higher than OTCs (p < 0.001 for each).

**Table 3A:** Matrix of pairwise comparisons for rescue medications across comfort variable.

|  | Triptan | Gepant | OTC | Ergot/Barb/Opioid |
| --- | --- | --- | --- | --- |
| Triptan | X | = | → | → |
| Gepant | = | X | → | → |
| OTC | ← | ← | X | = |
| Ergot/Barb/Opioid | ← | ← | = | X |

Each row indicates the reference group. → indicates row treatment significantly better than column treatment. ← indicates row treatment significantly worse than column treatment. = indicates no significant difference. X indicates same treatment (diagonal). Ergot/Barb/Opioid indicates the combined group of those on either ergot, barbiturate or opioid monotherapy.

#### Control

Participants using gepants reported greater perceived control than those using ergots/barbiturates/opioids (*p* = 0.031), triptans (*p* = 0.039), or OTCs (*p* < 0.001; Table 3B). Triptans were also rated higher than OTCs (*p* < 0.001), while differences between triptans and ergots/barbiturates/opioids (*p* = 0.105), and OTCs vs. ergots/barbiturates/opioids (*p* = 0.216), were not statistically significant.

**Table 3B:** Matrix of pairwise comparisons for rescue medications across control variable.

|  | Triptan | Gepant | OTC | Ergot/Barb/Opioid |
| --- | --- | --- | --- | --- |
| Triptan | X | ← | → | = |
| Gepant | → | X | → | → |
| OTC | ← | ← | X | = |
| Ergot/Barb/Opioid | = | ← | = | X |

Each row indicates the reference group. → indicates row treatment significantly better than column treatment. ← indicates row treatment significantly worse than column treatment. = indicates no significant difference. X indicates same treatment (diagonal). Ergot/Barb/Opioid indicates the combined group of those on either ergot, barbiturate or opioid monotherapy.

### Comfort and Control by Preventive Medication Class

Among the 1751 participants on preventive monotherapy, 281 (16.0%) were taking an anti-depressant, 307 (15.5%) an anti-seizure medication, 236 (13.5%) a beta blocker, 171 (9.8%) on OnabotulinumtoxinA, 235 (13.4%) on a gepant, and 487 (27.8%) on a CGRP antibody. Thirty-four participants who indicated “unsure” were excluded from this analysis.

#### Comfort

Participants taking antidepressants reported higher comfort than those on onabotulinumtoxinA (*p* = 0.017) but did not differ significantly from other groups (*p’*s > 0.081; Table 4A). Similarly, anti-seizure medications were associated with higher comfort than onabotulinumtoxinA (*p* = 0.015), but not with other classes (*p*s > 0.074). Gepants were rated more favorably than beta blockers (*p* = 0.011) and onabotulinumtoxinA (*p* = 0.002), and CGRP antibodies were associated with greater comfort than onabotulinumtoxinA (*p* = 0.012). Comfort ratings did not differ between onabotulinumtoxinA and beta blockers (*p* = 0.443).

**Table 4A:**
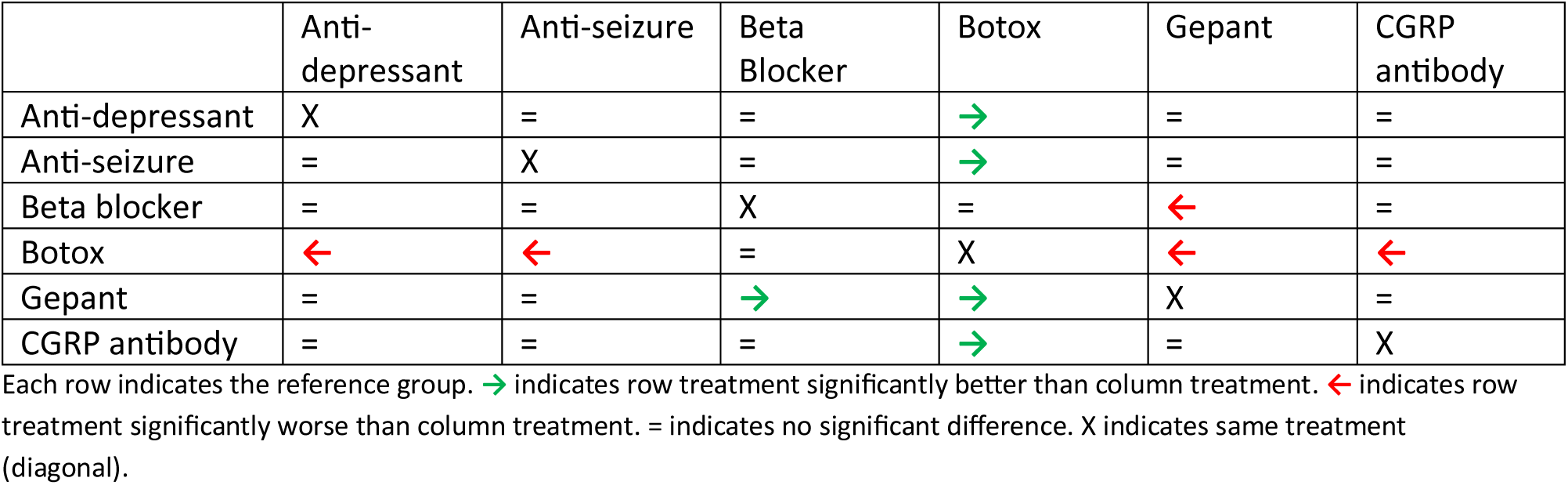
Matrix of pairwise comparisons for preventive medications across comfort variable.

#### Control

Antidepressants were associated with greater perceived control compared to onabotulinumtoxinA (*p* = 0.030), but not other groups (*p*s > 0.051; Table 4B). Anti-seizure medications were associated with higher control than beta blockers (*p* = 0.005) and onabotulinumtoxinA (*p* = 0.003). Gepants outperformed both beta blockers and onabotulinumtoxinA (*p* = 0.001 for both), as did CGRP antibodies (*p* = 0.020 and *p* = 0.013, respectively). There was no significant difference in perceived control between beta blockers and onabotulinumtoxinA (*p* = 0.679).

**Table 4B:** Matrix of pairwise comparisons for preventive medications across control variable.

|  | Anti-depressant | Anti-seizure | Beta Blocker | Botox | Gepant | CGRP antibody |
| --- | --- | --- | --- | --- | --- | --- |
| Anti-depressant | X | = | = | → | = | = |
| Anti-seizure | = | X | → | → | = | = |
| Beta blocker | = | ← | X | = | ← | ← |
| Botox | ← | ← | = | X | ← | ← |
| Gepant | = | = | → | → | X | = |
| CGRP antibody | = | = | → | → | = | X |

Each row indicates the reference group. → indicates row treatment significantly better than column treatment. ← indicates row treatment significantly worse than column treatment. = indicates no significant difference. X indicates same treatment (diagonal).

## Discussion

We identify patient and disease characteristics associated with perceived comfort with medications and control over symptoms in adults with migraine. Both measures were strongly related to mood symptoms and migraine-related disability, in addition to traditional clinical variables such as headache frequency and symptom severity. Importantly, only about one-quarter of participants reported high control despite nearly 60% reporting high comfort, underscoring that these are distinct constructs with different clinical implications. Furthermore, we identified that higher comfort and perceived efficacy rates in patients taking triptans and gepants amongst acute medication options, and lower comfort and efficacy in those taking beta blockers or OnabotulinumtoxinA amongst preventive medication options.

Higher headache frequency and symptom severity were associated with lower perceived comfort and control. While this is consistent with previously published studies, ^8,12^ the magnitude of this association in our analyses was modest, suggesting that clinical severity alone does not explain patients’ perceptions. In contrast, mood symptomatology and migraine-associated disability were much stronger predictors of perception of both comfort and control measures. Prior research has demonstrated bidirectional associations between migraine and depression or anxiety, and our results extend this work by showing that mood symptoms may outweigh attack frequency or severity in shaping perceptions of treatment adequacy and control.^21,33–36^ Clinically, this highlights the importance of routine screening for anxiety and depression in migraine care, as addressing mood symptoms may directly improve patients’ sense of treatment success.

Perception of disability is shaped not only by the frequency and intensity of migraine attacks but also by psychological factors such as mood and coping strategies.^34,37^ Higher perceived disability is linked to greater emotional distress, reduced participation in social and occupational roles, and lower overall well-being.^38,39^ Mood disturbances can exacerbate the perception of pain and reduce coping capacity, while migraines can, in turn, contribute to emotional distress, creating a reinforcing cycle between mood and migraine severity. Enhancing perceived comfort and control may be a key target in reducing the overall burden of migraine. Psychological interventions such as Acceptance and Commitment Therapy and cognitive-behavioral therapy have shown efficacy in migraine populations and may address both self-efficacy and treatment adherence.^40–42^

Mechanistically, brain areas conveying pain control have been shown to overlap with areas involved in migraine and mood disorders. For instance, prefrontal cortex activity has been shown to mediate pain control.^43,44^ Prefrontal activity may in turn mitigate activation of the amygdala and insula, which are areas with well-established roles in pain, migraine and mood disorders.^45–47^ Shared neurotransmitter systems, including serotonergic, dopaminergic, and PACAP pathways, further suggest biological overlap that could explain why patients with comorbid mood symptoms report lower comfort and control.^48,49^ These neurobiological intersections reinforce the need for integrated treatment approaches that address both migraine and psychiatric comorbidity.

Several demographic features were associated with comfort and control. Lower income and unemployment were related to both lower comfort and control while lower education was related only to lower comfort. Prior studies have reported an association between lower income and education with worse migraine burden, our study suggests areas of possible intervention, highlighting subgroups of patients who may need tailored education and support to improve treatment engagement and outcomes..^50–52^ Employment status also plays a role, as being employed can provide structure, insurance coverage, and financial stability, which may positively influence disease perception, while unemployment may exacerbate stress and feelings of vulnerability.^53,54^ Similar to these previous studies, our current data does not prove directionality of the relationships between these factors. These social determinants of health highlight subgroups of patients who may need tailored education and support to improve treatment engagement and outcomes. Age was also found to be significant for both comfort and control measures. However, effect sizes were very small and unlikely to be clinically meaningful, suggesting that age should not be a focus in interpreting patient perceptions.

We found that patients using a single acute or preventive medication reported the highest comfort and control, compared with those using no medications or multiple agents. For acute therapies, both gepants and triptans rated more favorably than OTC agents or opioids/barbiturates for comfort, while gepants were uniquely associated with the highest control ratings. There may be several explanations. These findings likely reflect the favorable efficacy and tolerability of migraine-specific medications (gepants and triptans) compared with less targeted or more side-effect-prone options.^55,56^ The absence of difference between triptans and gepants for comfort, but superiority of gepants for control, suggests that patients may view gepants as providing a stronger sense of reliability or predictability in attack management. Gepants also have relatively low side effect profiles, which may further contribute to higher reported comfort.^57^ Interestingly, despite evidence that NSAIDs and OTCs can be more effective than opioids or ergots in migraine^58,59^, our cohort did not perceive OTCs as superior. One explanation may be that NSAIDs are less effective in individuals with higher disease severity, a group overrepresented in our sample, leading to lower comfort and control ratings for OTCs in this study.^60^

Amongst preventive medications, onabotulinumtoxinA was consistently associated with lower comfort and control compared to other classes. This could reflect its use primarily in chronic migraine, a more severe population, though perception of limited benefit may also contribute.^61,62^ Additionally, the need for frequent clinician visits in order to administer onabotulinumtoxinA may be perceived by patients as an indicator of severity. Beta blockers were also associated with lower perceptions, despite evidence of effectiveness, suggesting a potential mismatch between clinical efficacy and patient experience.^63,64^ In contrast, gepants and CGRP monoclonal antibodies were viewed more favorably, echoing emerging real-world data showing high patient satisfaction with these novel agents. These differences emphasize the importance of considering not only efficacy and safety but also patient perceptions when selecting preventive treatments.

Having an available medication may provide individuals with a sense of control over their symptoms, while needing multiple agents may be perceived as a marker of refractory disease. Although polytherapy is often necessary and evidence supports multimodal approaches to migraine management,^65–67^ patients may interpret multiple medications as a sign of treatment failure. Clinicians should therefore proactively frame combination therapy as a strategy for optimizing care, rather than as a marker of poor prognosis, to maintain patient confidence and adherence.

### Limitations and Future Directions

The cross-sectional study design limits understanding of directionality. Future longitudinal studies may be provide insight on the longer term relationships between migraine symptomatology, perceived disease control, and perceived disability over time. The sample also represents a fairly severe cohort, with over 80% of individuals reporting severe disability per MIDAS score, as thus may not be generalizable to less severe populations. The cohort was largely white and of high education level, thus highlighting the need for future studies in diverse patient cohorts. Also, calcium channel blockers and angiotensin receptor blockers were not accounted for in data collection or analysis. Future longitudinal work should examine how comfort and control evolve over time with changes in mood, disability, and treatment strategies, and whether interventions targeting self-efficacy or addressing socioeconomic barriers can improve both perceptions and clinical outcomes.

## Conclusion

In conclusion, perceived comfort with migraine medications and control of migraine symptoms were most strongly associated with mood symptoms and disability, rather than headache frequency or severity. Socioeconomic factors such as income, education, and employment further shaped these perceptions. Patients on triptans and gepants reported more favorable perceptions amongst acute medication classes, while those on beta blockers and onabotulinumtoxinA reported less favorable opinions compared to other preventive classes.

These findings suggest that clinicians should integrate patient perceptions into treatment planning, particularly by addressing mood symptoms, clarifying expectations for preventive therapies, and tailoring communication for vulnerable populations. Doing so may improve adherence, satisfaction, and ultimately real-world outcomes in migraine care.

## Data Availability

All data produced in the present study are available upon reasonable request to the authors

https://faculty.sites.uci.edu/neuroinformatics/head-us/

## Abbreviations

PRO: patient reported outcome
HeAD-US: Headache Assessment via Digital Platform in United States
CIRBI: Center for IRB Intelligence
IRB: institutional review board
ICHD-3: International Classification of Headache Disorders ‐ 3
AMS: American Migraine Study
AMPP: American Migraine Prevalence and Prevention
mTOQ: Migraine Treatment Optimization Questionnaire ‐ 6
MSSS: Migraine Symptom Severity Score
MIDAS: Migraine Disability Assessment Score
PHQ-4: Patient Health Questionnaire-4
OTC: Over the counter
CGRP: calcitonin gene related peptide
SPSS: IBM Statistical Package for Social Science

## Notes

### Competing Interest Statement

Kevin M. Spiegler reports no conflicts of interest.
Angeliki Vgontzas reports no conflicts of interest.
Kristina M Fanning is the managing director of MIST Research LLC which received grants from the National Headache Foundation in addition to funding from Allergan, Amgen, Dr. Reddy Laboratories/Promius, and Eli Lilly via collaboration with Vedanta Research.
Ryan Bostic is an employee of MIST research.
Alexandre Urani is an employee of APTAR LLC, the parent company of Migraine Buddy.
Francois Cadiou is a former employee of APTAR LLC, the parent company of Migraine Buddy.
Richard B. Lipton receives research support from the NIH and the FDA as well as the National Headache Foundation and the Marx Foundation. He serves on the editorial board of Neurology, senior advisor to Headache, and associate editor to Cephalalgia. He has reviewed for the NIA and NINDS, holds stock options in Axon, Biohaven Holdings, CoolTech and Manistee; serves as consultant, advisory board member, or has received honoraria from: Abbvie (Allergan), American Academy of Neurology, American Headache Society, Amgen, Avanir, Biohaven, Biovision, Boston Scientific, CoolTech, Dr. Reddy (Promius), Electrocore, Eli Lilly, eNeura Therapeutics, GlaxoSmithKline, Grifols, Lundbeck (Alder), Pfizer, Teva, Trigemina, Vector, Vedanta. He receives royalties from Wolffs Headache 7th and 8th Edition, Oxford Press University, 2009, Wiley and Informa.
Ali Ezzati receives research support from the following sources: National Institute of Health (K23 AG063993; R01 AG080635; R01 AG095017); the Alzheimer Association (SG 24 988292), Cure Alzheimer Fund, and Amgen investigator initiated studies.

### Funding Statement

This study did not receive any funding.

### Author Declarations

IRB of University of California Irvine gave ethical approval for this work. (CIRBI protocol # Pro00072897)

IRB of Brigham and Womens Hospital gave ethical approval for this work (IRB # 2025P000071).

