## Supplemental Tables for "Perceived comfort with migraine treatments and control of migraine symptoms: results from the HEAD-US study"

Supplemental Table 1A: Outcome variables presented numerically for comfort variable

| Outcome Variable | Low Comfort (n =2372) | High Comfort (n = 3345) | p value |
|---|---|---|---|
| Monthly Headache Days, *avg (SD)* | 14.19 (8.57) | 9.88 (7.28) | **< 0.001** |
| MSSS, *(median, IQR)* | 19 [17-20] | 18 [16-20] | **< 0.001** |
| MIDAS | | | **< 0.001** |
| Little to None | 22 (0.93) | 157 (4.69) | *** |
| Mild | 44 (1.86) | 240 (7.17) | *** |
| Moderate | 144 (6.07) | 517 (15.46) | *** |
| Severe | 2162 (91.15) | 2431 (72.68) | *** |
| PHQ-4 | | | **< 0.001** |
| Normal | 550 (23.19) | 1427 (42.89) | *** |
| Mild | 757 (31.91) | 1046 (32.27) | |
| Moderate | 566 (23.86) | 553 (16.53) | *** |
| Severe | 499 (21.04) | 319 (9.54) | *** |

Data are presented as total number (percentage) unless otherwise specified. Comparisons reaching statistical significance ($p < 0.05$) are bolded. *** denotes significance after post hoc testing. Avg = average, SD = standard deviation, IQR = interquartile range.

Supplemental Table 1B: Outcome variables presented numerically for control variable

| Outcome Variable | Low Control (n =4142) | High Control (n = 1575) | p value |
|---|---|---|---|
| Monthly Headache Days, *avg (SD)* | 12.68 (8.36) | 8.99 (6.78) | **< 0.001** |
| MSSS, *(median, IQR)* | 19 [17-20] | 18 [16-19] | **< 0.001** |
| MIDAS | | | **< 0.001** |
| Little to None | 73 (1.76) | 106 (6.73) | *** |
| Mild | 133 (3.21) | 151 (9.59) | *** |
| Moderate | 383 (9.25) | 278 (17.65) | *** |
| Severe | 3553 (85.78) | 1040 (66.03) | *** |
| PHQ | | | **< 0.001** |
| Normal | 1210 (29.21) | 767 (48.70) | *** |
| Mild | 1345 (32.47) | 458 (29.08) | |
| Moderate | 888 (21.44) | 230 (14.60) | *** |
| Severe | 698 (16.85) | 120 (7.62) | *** |

Data are presented as total number (percentage) unless otherwise specified. Comparisons reaching statistical significance ($p < 0.05$) are bolded. *** denotes significance after post hoc testing. Avg = average, SD = standard deviation, IQR = interquartile range.

Supplemental Table 2A: Medication use in patients on monotherapy for comfort variable

| Medication | Total, *n* | High Comfort, *n (%)* | Low Comfort, *n (%)* |
|---|---|---|---|
| Acute | | | |
| Over-The-Counter | 1018 | 536 (52.7) | 482 (47.3) |
| Gepant | 570 | 375 (65.8) | 195 (34.2) |
| Triptan | 727 | 471 (64.7) | 256 (35.3) |
| Barbiturate/Opioid/Ergot | 105 | 53 (50.5) | 52 (49.5) |
| Preventive | | | |
| CGRP monoclonal antibody | 487 | 315 (64.7) | 172 (35.3) |
| Anti-depressant | 281 | 183 (65.2) | 98 (34.8) |
| Anti-seizure | 307 | 200 (65.1) | 107 (34.9) |
| Beta Blocker | 236 | 136 (57.6) | 100 (42.4) |
| OnabotulinumtoxinA | 171 | 92 (53.8) | 79 (46.2) |
| Gepant | 235 | 162 (69.0) | 73 (31.0%) |

Supplemental Table 2B: Medication use in patients on monotherapy for control variable

| Medication | Total, *n* | High Control, *n (%)* | Low Control, *n (%)* |
|---|---|---|---|
| Acute | | | |
| Over-The-Counter | 1018 | 210 (20.6) | 808 (79.4) |
| Gepant | 570 | 221 (38.8) | 349 (61.2) |
| Triptan | 727 | 244 (33.6) | 483 (66.4) |
| Barbiturate/Opioid/Ergot | 105 | 29 (27.6) | 76 (72.4) |
| Preventive | | | |
| CGRP monoclonal antibody | 487 | 161 (33.1) | 326 (66.9) |
| Anti-depressant | 281 | 91 (32.4) | 190 (67.6) |
| Anti-seizure | 307 | 110 (35.8) | 197 (64.2) |
| Beta Blocker | 236 | 58 (24.6) | 178 (75.4) |
| OnabotulinumtoxinA | 171 | 39 (22.8) | 132 (77.2) |
| Gepant | 235 | 90 (38.3) | 145 (61.7) |
